# Health Care Workers Knowledge, Attitudes, and Practice Towards Childhood Tuberculosis in Primary Health Facilities in Lusaka, Zambia

**DOI:** 10.1101/2023.06.16.23291512

**Authors:** Paul Chabala Kaumba, Mary Kagujje, Chalilwe Chungu, Sarah Nyangu, Nsala Sanjase, Minyoi Mubita Maimbolwa, Brian Shuma, Lophina Chilukutu, Monde Muyoyeta

## Abstract

**BACKGROUND:** Zambia is among the countries with high tuberculosis (TB) and Tuberculosis/Human Immunodeficiency Virus (TB/HIV) burdens, with a significant number of cases affecting children. However, a considerable portion of TB cases, especially in children, remains undiagnosed. This study aimed to assess and understand the factors influencing the knowledge, attitudes, and practices of health care workers (HCWs) regarding childhood TB in Zambia.

**METHOD:** Using a cross-sectional survey design, a self-administered anonymous questionnaire was employed to evaluate HCWs’ knowledge, attitudes, and practices related to TB. The study was conducted at two primary health facilities in Lusaka, Zambia, between July and August 2020. The questionnaire data collected was later transcribed to an electronic system called DHIS 2.

**RESULTS:** Out of 238 participants, the majority (72.3%) were female HCWs. Most staff members across various departments demonstrated awareness of the primary TB symptom, which is coughing. However, approximately half of the participants had limited knowledge regarding the transmission of TB through oral activities such as singing and laughing. Furthermore, the study found that 21.4% of HCWs reported regular interaction with children in their daily work, while 17.2% did not.

Among the HCWs, 73.1% referred children to the TB clinic to submit a sputum sample, 55.5% requested a sample from the patient, 55.9% expedited the process for children, 58.4% provided education on cough etiquette, and 42.4% recorded the child in the presumptive TB register. Only a negligible 0.8% of HCWs did not take any action for children exhibiting TB symptoms.

**CONCLUSION:** The study highlighted variations in knowledge levels based on gender, department, and training history among the HCWs. Those working in the TB department generally exhibited better knowledge and attitudes regarding TB, with a 50/50% variation. Therefore, it is crucial to enhance the TB knowledge, attitudes, and practices of female HCWs, who constitute most staff involved in TB diagnosis and treatment.

This study emphasizes the importance of improving the understanding of childhood TB among HCWs, particularly among female staff. Enhancing their knowledge and attitudes towards TB will contribute to early diagnosis and improved management of TB cases, ultimately reducing the burden of childhood TB in Zambia.

## Introduction

Globally, an estimated 6,431,705 people developed tuberculosis in 2021 of which about 7% were children (1). Majority (about 96%) of children who died from TB were not on treatment (2). According to the World Health Organization (WHO), at least 10% of all TB notifications in high burden countries should be made to children under the age of 15(1). However, most high burden countries did not meet this goal, Zambia included. In the three years leading up to 2022, Zambia reported approximately 40,000 annual cases of TB, with less than 10% of those cases being in children (1). According to estimates, the country missed 59 000 cases of tuberculosis in 2021, of which 5,500 were in children. (3)

Due to the pauci-bacillary nature of pediatric TB and its unique presentation in children, pediatric TB diagnosis can be difficult. (4–6) Variable knowledge, attitudes, and practices (KAP) concerning childhood TB among healthcare professionals are among the factors influencing the ineffective case detection of pediatric tuberculosis. In other settings, KAP studies on pediatric TB found that HCWs continue to hold stigmatizing beliefs and have misconceptions about TB diagnosis, treatment, and infection prevention (7–12). Even in light of new TB training, overall knowledge of the disease is insufficient (3, 5).

Children with symptoms of TB commonly present to primary healthcare facilities but due to limited capacity to recognize presumptive TB in children and due to the complexities of diagnosing childhood TB, (13) they are often referred from primary health facilities to referral hospitals and specialized pediatric units resulting in delayed diagnosis.(14) Optimizing the knowledge, attitudes, and practices (KAP) of health care workers, especially at primary health care level, is critical to early diagnosis and treatment of childhood TB.(15)

As far as we know, there is currently no local literature on KAP among health workers towards childhood TB in Zambia. We undertook a KAP study to identify potential health system barriers to childhood TB case detection. This data is intended to inform programmatic decision making on priority interventions towards improvement of childhood TB case detection.

## Methods

### Study design and participants

We conducted a cross-sectional study at two high volume primary health facilities in Lusaka, Zambia, Kanyama, and Chawama, between July 2020 and August 2020. Registered health care workers (nurse, clinical officer and medical officer) working at the health facility participated in the study. The main study population was made up of children between the ages 0-14 presenting to intervention health facilities and house contacts of TB patients notified at Kanyama and Chawama. Chipata and Matero health facilities were Control Sites. Only TB case detection trends were observed from the control sites. The intervention sites are on average 14km away from the control sites and this made for the expectation to minimise the risk of contamination.

### Data collection and study tools

A paper-based structured questionnaire with 37 questions, adapted from multiple sources but customized to the Zambian setting, was self-administered by the health care workers (16). The Knowledge, Attitudes and Practices (KAP), questionnaire contained three sections divided into Knowledge, Attitudes and Practices. The questionnaire included questions on general demographic information including age, gender, profession, department, duration at facility, TB training history and length of training, knowledge of the participant, HCWs attitude and practices towards TB. Additionally, the questionnaire had 4 questions on TB awareness and sources of information.

Completed paper-based questionnaires were securely kept at the health facility with access limited to the study coordinators and the facility in-charge. The questionnaires were then captured into the DHIS 2-based electronic database by qualified research staff. The data extracted from DHIS 2 was secured under password-protected access systems identified by coded ID numbers only. Only de-identified data was stored on a secure server managed by CIDRZ.

### Statistical analysis

All questions on knowledge were assigned an overall score of one point. Where a question had only one correct response, each correct answer was assigned one point and zero points were assigned for both incorrect and ‘don’t know/not sure’ answer. When a question had more than one correct response, each correct response contributed equal weight towards the overall score and each incorrect response carried no weight towards the overall score for the question. The points for each question were added together to create a composite knowledge score, ranging from 0 to 14. All Results of quantitative variables were reported as frequency (percentage %).

Descriptive analysis was done to describe the population and summarize the responses for sociodemographic information and for questions that had free text required of the respondents as part of the answers. The results were presented as frequencies and percentages; chi-squared test and Fisher’s exact test were used to compare groups as appropriate.

### Ethical considerations

Approval for study implementation was obtained from the University of Zambia Biomedical Research Ethical Committee (UNZABREC), IRB Reference number 635-2020 under the TB REACH wave 7 study. The respondents provided written informed consent before being allowed to complete the paper based self-administered questionnaire. The survey protected the confidentiality of the respondents by maintaining anonymous responses.

## Results

A total of 238 participants completed the surveys (giving a 100% response rate), and all 238 were included in the analysis. The majority 151 (63.4%) were less than 30 years of age, 66 (27.7%) were male, and 93 (39.1%) were from the Outpatient department (OPD) (Table 1). Most participants, 128 (53.8%), had been in employment for more than 1 year and less than half of the staff, 100 (42.0%) reported previous training in TB. Of those who had been previously trained, only 35 (35.0%) had been trained specifically on childhood TB. Sixty one percent (61%) of those trained had been trained within 1 year of conducting the survey.

**Table 1:** Sociodemographic characteristics of HCWs

| **Variables** | **Frequency (%)** |
| --- | --- |
| **Age group** |  |
| Under 30 | 151 (63.4) |
| 31-40 | 61 (25.6) |
| 41-50 | 17 (7.1) |
| Over 50 | 7 (2.9) |
| Unknown | 2 (0.8) |
| **Sex** |  |
| Male | 66 (27.7) |
| Female | 172 (72.3) |
| **Department** |  |
| Outpatient dept. | 93 (39.1) |
| ART | 27 (11.3) |
| IPD | 74 (31.1) |
| MCH | 38 (16.0) |
| TB | 4 (1.7) |
| VMMC | 1 (0.4) |
| Unknown | 1 (0.4) |
| **Duration employed** |  |
| Less than 1 year | 110 (46.2) |
| Between 1-3 years | 92 (38.7) |
| Between 3-5 years | 12 (5.0) |
| Between 5-10 years | 15 (6.3) |
| More than 10 years | 9 (3.8) |
| **Training** |  |
| Ever trained on TB | 100 (42.0) |
| Adult TB | 47 (47.0) |
| Childhood TB | 35 (35.0) |
| TPT | 40 (40.0) |
| Infection Control | 29 (29.3) |
| Case Finding | 12 (12.0) |
| TB HIV | 53 (53.0) |
| **How long ago were you trained?** |  |
| < 3 months | 25 (25.0) |
| ≥3 months - < 6 months | 18 (18.0) |
| Less than 1 year (≥6 months-≤1 year) | 18 (18.0) |
| ≥1 year to 3 years | 12 (12.0) |
| ≥ 3 years | 27 (27.0) |

Most of the staff were aware of the major TB symptoms (98.9%), with about half of the staff showing little knowledge around the oral activities that can cause TB transmission; only 52.7% of the OPD staff knew about the oral activity’s (singing and laughing) ability to transmit TB. All the staff responded that coughing spreads TB except 1 staff from OPD(Table 2). All the respondents said that TB affects the lungs while 52.7% from OPD responded that it can also affect the heart. All the TB clinic staff responded favorably to TB’s ability to affect the lungs, heart, spine, meninges, lymph nodes, abdomen and pleura while 2 (50%) responded favorably to TB’s ability to affect the larynx. The VMMC staff responded that lack of vaccination, HIV infection, malnutrition and contact to TB patients are among the childhood TB infection risk factors. However, they unfavorably responded to ‘being under 5 and a resident in Zambia,’ which are also TB risk factors.

**Table 2:** Knowledge of HCWs on Tuberculosis

| Variable | Correct response (%) |  |  |  |  |  |  |
| --- | --- | --- | --- | --- | --- | --- | --- |
|  | Outpatient dept. (N=93) | ART (N=27) | IPD (N=74) | MCH (N=38) | TB (N=4) | VMMC (N=1) | Unknown (N=1) |
| **Zambia is high burden TB** |  |  |  |  |  |  |  |
| Yes | 92 (98.9) | 27 (100.0) | 74 (100.0) | 38 (100.0) | 4 (100.0) | 1 (100.0) | 1 (100.0) |
| **How is TB spread? (Coughing)** |  |  |  |  |  |  |  |
| Coughing | 92 (98.9) | 27 (100.0) | 74 (100.0) | 38 (100.0) | 4 (100.0) | 1 (100.0) | 1 (100.0) |
| Sneezing | 74 (79.6) | 19 (70.4) | 46 (62.2) | 25 (65.8) | 2 (50.0) | 1 (100.0) | 1 (100.0) |
| Singing | 49 (52.7) | 7 (25.9) | 18 (24.3) | 14 (36.8) | 2 (50.0) | 0 (0.0) | 0 (0.0) |
| Laughing | 49 (52.7) | 6 (22.2) | 20 (27.0) | 12 (31.6) | 2 (50.0) | 0 (0.0) | 0 (0.0) |
| Skin contact | 84 (90.3) | 26 (96.3) | 73 (98.6) | 34 (89.5) | 4 (100.0) | 1 (100.0) | 1 (100.0) |
| **TB can affect the following body parts?** |  |  |  |  |  |  |  |
| Lungs | 93 (100.0) | 27 (100.0) | 74 (100.0) | 38 (100.0) | 4 (100.0) | 1 (100.0) | 1 (100.0) |
| Larynx | 55 (59.1) | 12 (44.4) | 30 (40.5) | 19 (50.0) | 2 (50.0) | 1 (100.0) | 0 (0.0) |
| Heart | 49 (52.7) | 14 (51.9) | 21 (28.4) | 12 (31.6) | 4 (100.0) | 0 (0.0) | 0 (0.0) |
| Spine | 88 (94.6) | 24 (88.9) | 68 (91.9) | 34 (89.5) | 4 (100.0) | 1 (100.0) | 1 (100.0) |
| Meninges | 77 (82.8) | 20 (74.1) | 63 (85.1) | 30 (78.9) | 4 (100.0) | 1 (100.0) | 1 (100.0) |
| Lymph nodes | 69 (74.2) | 20 (74.1) | 51 (68.9) | 23 (60.5) | 4 (100.0) | 1 (100.0) | 1 (100.0) |
| Abdomen | 80 (86.0) | 23 (85.2) | 64 (86.5) | 27 (71.1) | 4 (100.0) | 1 (100.0) | 1 (100.0) |
| Pleura | 71 (76.3) | 17 (63.0) | 42 (56.8) | 20 (52.6) | 4 (100.0) | 1 (100.0) | 0 (0.0) |
| **Risk factor for childhood TB infection** |  |  |  |  |  |  |  |
| Not being vaccinated with BCG | 82 (88.2) | 24 (88.9) | 68 (91.9) | 35 (92.1) | 1 (25.0) | 1 (100.0) | 1 (100.0) |
| HIV | 80 (86.0) | 24 (88.9) | 64 (86.5) | 35 (92.1) | 3 (75.0) | 1 (100.0) | 1 (100.0) |
| Malnutrition | 75 (80.6) | 23 (85.2) | 59 (79.7) | 35 (92.1) | 3 (75.0) | 1 (100.0) | 1 (100.0) |
| Being less than 5 years old | 58 (62.4) | 15 (55.6) | 38 (51.4) | 17 (44.7) | 2 (50.0) | 1 (100.0) | 0 (0.0) |
| Living in Zambia | 31 (33.3) | 3 (11.1) | 25 (33.8) | 9 (23.7) | 2 (50.0) | 0 (0.0) | 0 (0.0) |
| Being a contact to a TB case | 79 (84.9) | 26 (96.3) | 60 (81.1) | 34 (89.5) | 3 (75.0) | 1 (100.0) | 1 (100.0) |

Table 3 shows that 51 (21.4%) of the HCWs spent every day with children while 41(17.2%) indicated to have no contact with children. Of the 238 HCWs, 174 (73.1%) refer the children to the TB clinic to submit a sputum sample. Additionally, 132 (55.5%) request the patient for a sample, 133 (55.9%) fast track the children, 139 (58.4%) provide education on cough etiquette and 101 (42.4%) record the child in the presumptive TB register. Only 2 (0.8%) of the HCWs said they do nothing about children with TB symptoms. Most of the HCW responded that they use GeneXpert as a reliable test to diagnose TB in Children while 11 (4.6%) indicated no knowledge on the test to use. Most HCWs 187 (78.6%) indicated they use gastric aspirates to collect sputum samples.

**Table 3:** Practices of HCWs on Childhood TB

| Practice | Frequency n=238 (%) |
| --- | --- |
| **How often do you interact with children with symptoms suggestive of TB?** |  |
| Everyday | 51 (21.4) |
| More than once a week | 42 (17.6) |
| At least once a week | 49 (20.6) |
| Once a month | 48 (20.2) |
| Never | 41 (17.2) |
| Unknown | 7 (2.9) |
| **What do you do when you identify a child with symptoms of TB? (True responses)** |  |
| Ask the patient to go to TB corner to submit a sputum sample | 174 (73.1) |
| Request the patient to submit a sputum sample | 132 (55.5) |
| Fast track the child | 133 (55.9) |
| Provide education on cough etiquette | 139 (58.4) |
| Document the child in the presumptive TB register | 101 (42.4) |
| Nothing | 2 (0.8) |
| **Which test do you often rely on to diagnose TB in children?** |  |
| Sputum microscopy | 39 (16.4) |
| Gene Xpert | 114 (47.9) |
| Chest X-ray | 47 (19.7) |
| LAM | 12 (5.0) |
| Culture and DST | 12 (5.0) |
| Other | 2 (0.8) |
| N/A | 1 (0.4) |
| Unknown | 11 (4.6) |
| **What do you do for children not able to produce sputum?** |  |
| Prescribe antibiotics for 1 week and then ask them to return to the health facility for review | 18 (7.6) |
| Collect gastric aspirates | 187 (78.6) |
| Use chest x-ray to make a diagnosis | 122 (51.3) |
| Give mother sputum bottle to continue trying to get sputum from the child | 36 (15.1) |
| Request for LAM | 52 (21.8) |
| Other | 5 (2.1) |
| N/A | 1 (0.4) |

The results further showed a strong association between TB knowledge and the department where staff worked.

Table 4: HCWs characteristics and knowledge level on TB Infection Control below shows the characteristics and knowledge levels of healthcare workers on TB infection control. ART department staff had lower knowledge of TB, evidenced by 17 (63%) respondents giving incorrect answers to questions about TB infection control, versus 1 (3.7%) respondent who gave correct answers, and 9 (33.3%) respondents who did not respond to the questions about infection control. In the TB clinic, 2 (50%) respondents gave the correct responses, while 2(50%) gave the incorrect ones. Additionally, the OPD/IPD HCWs had 42 (25.1%) wrong responses compared to 21 (12.6%) correct responses with majority 104 (62.3%) not giving responses to the questions. The findings showed that there was no association between TB infection control knowledge, Age and Sex. However, there was a close association (p=0.07) between TB infection control knowledge and the duration of employment.

**Table 4:** HCWs characteristics and knowledge level on TB Infection Control

| HCWs' characteristics | TB Infection Control Knowledge level |  |  | p-value |
| --- | --- | --- | --- | --- |
|  | Bad (n=70) | Good (n=29) | Unknown (n=139) |  |
| **Age group** |  |  |  |  |
| Under 30 | 40 (26.5) | 18 (11.9) | 93 (61.6) | 0.21 |
| 31-40 | 23 (37.7) | 5 (8.2) | 33 (54.1) |  |
| 41-50 | 6 (35.3) | 3 (17.6) | 8 (47.1) |  |
| Over 50 | 1 (14.3) | 3 (42.9) | 3 (42.9) |  |
| Unknown | 0 (0.0) | 0 (0.0) | 2 (100.0) |  |
| **Age (Median, IQR) (Under 30, Under 30 to 31-40)** |  |  |  |  |
| **Sex** |  |  |  |  |
| Male | 25 (37.9) | 9 (13.6) | 32 (48.5) | 0.14 |
| Female | 45 (26.2) | 20 (11.6) | 107 (62.2) |  |
| **Department** |  |  |  |  |
| Outpatient dept./IPD | 42 (25.1) | 21 (12.6) | 104 (62.3) | 0.00* |
| ART | 17 (63.0) | 1 (3.7) | 9 (33.3) |  |
| MCH | 9 (23.7) | 4 (10.5) | 25 (65.8) |  |
| TB | 2 (50.0) | 2 (50.0) | 0 (0.0) |  |
| VMMC | 0 (0.0) | 0 (0.0) | 1 (100.0) |  |
| Unknown | 0 (0.0) | 1 (100.0) | 0 (0.0) |  |
| **Duration employed** |  |  |  |  |
| Less than 1 year | 25 (22.7) | 14 (12.7) | 71 (64.5) | 0.07 |
| Between 1-3 years | 34 (37.0) | 8 (8.7) | 50 (54.3) |  |
| Between 5-3 years | 4 (33.3) | 1 (8.3) | 7 (58.3) |  |
| Between 5-10 years | 4 (26.7) | 2 (13.3) | 9 (60.0) |  |
| More than 10 years | 3 (33.3) | 4 (44.4) | 2 (22.2) |  |

\* P value <0.05

### The HealthCare Worker’s KAP Scores

The characteristics and scores of healthcare workers in each category of the Knowledge, Attitudes, and Practices (KAP) survey are presented in Table 6: HCWs characteristics and knowledge score below and in Table 5: Detailed HCWs characteristics and knowledge score of the supplementary data. Knowledge was subdivided further into Epidemiology, Diagnosis, and Treatment. The results revealed that, older HCWs (Over 50 years old) scored the highest on the KAP for Knowledge (9.6; 68.6%), Attitude (2.4; 80%), and Practices (1.2; 60%) whereas the staff members who did not provide their Age group scored the lowest. Male participants scored higher than female participants: males scored knowledge 8.8 (62.9%), attitudes 2.6 (86.7%), practices 1.1 (55%) and females 8.3 (59.3%) knowledge, 2.3 (76.7%) attitudes, 0.8 (40%) practices.

**Table 5:** Detailed HCWs characteristics and knowledge score

| HCWs' characteristic variable | Knowledge |  |  |  |  |  | Knowledge Score |  | Attitudes |  | Practices |  | Total |  |
| --- | --- | --- | --- | --- | --- | --- | --- | --- | --- | --- | --- | --- | --- | --- |
|  | Epidemiology and transmission |  | Diagnosis |  | Treatment |  |  |  |  |  |  |  |  |  |
|  | Maximum Score=4 | % | Maximum score=2 | % | Maximum score=8 | % | Maximum Score=14 | % | Maximum Score=3 | % | Maximum score=2 | % | Maximum score=19 | % |
| Age group |  |  |  |  |  |  |  |  |  |  |  |  |  |  |
| Under 30 | 2.9 | 72.5% | 1 | 50.0% | 4.5 | 56.3% | 8.4 | 60.0% | 2.4 | 80.0% | 0.9 | 45.0% | 11.7 | 61.6% |
| 31-40 | 2.7 | 67.5% | 1.1 | 55.0% | 4.7 | 58.8% | 8.5 | 60.7% | 2.4 | 80.0% | 0.9 | 45.0% | 11.8 | 62.1% |
| 41-50 | 2.7 | 67.5% | 1.1 | 55.0% | 4.6 | 57.5% | 8.4 | 60.0% | 2.3 | 76.7% | 0.9 | 45.0% | 11.6 | 61.1% |
| Over 50 | 3.2 | 80.0% | 1.3 | 65.0% | 5.1 | 63.8% | 9.6 | 68.6% | 2.4 | 80.0% | 1.2 | 60.0% | 13.2 | 69.5% |
| Unknown | 3.2 | 80.0% | 1.2 | 60.0% | 3.5 | 43.8% | 7.9 | 56.4% | 2.3 | 76.7% | 0.5 | 25.0% | 10.7 | 56.3% |
| Sex |  |  |  |  |  |  |  |  |  |  |  |  |  |  |
| Male | 2.9 | 72.5% | 1.1 | 55.0% | 4.8 | 60.0% | 8.8 | 62.9% | 2.6 | 86.7% | 1.1 | 55.0% | 12.5 | 65.8% |
| Female | 2.8 | 70.0% | 1 | 50.0% | 4.5 | 56.3% | 8.3 | 59.3% | 2.3 | 76.7% | 0.8 | 40.0% | 11.4 | 60.0% |
| Department |  |  |  |  |  |  |  |  |  |  |  |  |  |  |
| Outpatient dept. | 3 | 75.0% | 1 | 50.0% | 4.7 | 58.8% | 8.7 | 62.1% | 2.5 | 83.3% | 0.8 | 40.0% | 12 | 63.2% |
| ART | 2.7 | 67.5% | 1.1 | 55.0% | 5.3 | 66.3% | 9.1 | 65.0% | 2.5 | 83.3% | 0.6 | 30.0% | 12.2 | 64.2% |
| IPD | 2.7 | 67.5% | 1 | 50.0% | 4.1 | 51.3% | 7.8 | 55.7% | 2.4 | 80.0% | 0.9 | 45.0% | 11.1 | 58.4% |
| MCH | 2.6 | 65.0% | 0.9 | 45.0% | 4.4 | 55.0% | 7.9 | 56.4% | 2.1 | 70.0% | 0.9 | 45.0% | 10.9 | 57.4% |
| TB | 3.2 | 80.0% | 1.7 | 85.0% | 6.7 | 83.8% | 11.6 | 82.9% | 2.6 | 86.7% | 1.7 | 85.0% | 15.9 | 83.7% |
| VMMC | 1.9 | 47.5% | 1 | 50.0% | 4.6 | 57.5% | 7.5 | 53.6% | 2.4 | 80.0% | 0.9 | 45.0% | 10.8 | 56.8% |
| Unknown | 2.7 | 67.5% | 1 | 50.0% | 3.8 | 47.5% | 7.5 | 53.6% | 2.7 | 90.0% | 1 | 50.0% | 11.2 | 58.9% |
| Duration employed |  |  |  |  |  |  |  |  |  |  |  |  |  |  |
| Less than 1 year | 2.7 | 67.5% | 1 | 50.0% | 4.5 | 56.3% | 8.2 | 58.6% | 2.3 | 76.7% | 0.8 | 40.0% | 11.3 | 59.5% |
| Between 1-3 years | 2.9 | 72.5% | 1.1 | 55.0% | 4.8 | 60.0% | 8.8 | 62.9% | 2.5 | 83.3% | 0.9 | 45.0% | 12.2 | 64.2% |
| Between 3-5 years | 2.6 | 65.0% | 1.1 | 55.0% | 4.6 | 57.5% | 8.3 | 59.3% | 2.4 | 80.0% | 1.1 | 55.0% | 11.8 | 62.1% |
| Between 5-10 years | 2.5 | 62.5% | 1.1 | 55.0% | 4.1 | 51.3% | 7.7 | 55.0% | 2.4 | 80.0% | 1.1 | 55.0% | 11.2 | 58.9% |
| More than 10 years | 2.9 | 72.5% | 0.9 | 45.0% | 4.9 | 61.3% | 8.7 | 62.1% | 2.3 | 76.7% | 1.1 | 55.0% | 12.1 | 63.7% |
| Unknown | 2.6 | 65.0% | 1 | 50.0% | 4.6 | 57.5% | 8.4 | 60.0% | 2.5 | 83.3% | 0.9 | 45.0% | 11.8 | 62.1% |
| Ever trained on TB |  |  |  |  |  |  |  |  |  |  |  |  |  |  |
| No | 2.7 | 67.5% | 1 | 50.0% | 4.2 | 52.5% | 7.9 | 56.4% | 2.3 | 76.7% | 0.8 | 40.0% | 11 | 57.9% |
| Yes | 2.9 | 72.5% | 1.1 | 55.0% | 5.1 | 63.8% | 9.1 | 65.0% | 2.5 | 83.3% | 1 | 50.0% | 12.6 | 66.3% |
| Unknown | 2.6 | 65.0% | 0.9 | 45.0% | 4.3 | 53.8% | 7.8 | 55.7% | 2.5 | 83.3% | 0.9 | 45.0% | 11.2 | 58.9% |
| **Childhood TB** |  | 0.0% |  |  |  |  |  |  |  |  |  |  |  |  |
| No | 2.9 | 72.5% | 1 | 50.0% | 4.9 | 61.3% | 8.8 | 62.9% | 2.5 | 83.3% | 0.9 | 45.0% | 12.2 | 64.2% |
| Yes | 3 | 75.0% | 1.4 | 70.0% | 5.4 | 67.5% | 9.8 | 70.0% | 2.4 | 80.0% | 1.1 | 55.0% | 13.3 | 70.0% |

**Table 6:** HCWs characteristics and knowledge score

| HCWs' characteristic variable | Knowledge Score |  | Attitudes |  | Practices |  | Total |  |
| --- | --- | --- | --- | --- | --- | --- | --- | --- |
|  | Maximum Score=14 | % | Maximum Score=3 | % | Maximum score=2 | % | Maximum score=19 | % |
| **Age group** |  |  |  |  |  |  |  |  |
| Under 30 | 8.4 | 60.0% | 2.4 | 80.0% | 0.9 | 45.0% | 11.7 | 61.6% |
| 31-40 | 8.5 | 60.7% | 2.4 | 80.0% | 0.9 | 45.0% | 11.8 | 62.1% |
| 41-50 | 8.4 | 60.0% | 2.3 | 76.7% | 0.9 | 45.0% | 11.6 | 61.1% |
| Over 50 | 9.6 | 68.6% | 2.4 | 80.0% | 1.2 | 60.0% | 13.2 | 69.5% |
| Unknown | 7.9 | 56.4% | 2.3 | 76.7% | 0.5 | 25.0% | 10.7 | 56.3% |
| **Sex** |  |  |  |  |  |  |  |  |
| Male | 8.8 | 62.9% | 2.6 | 86.7% | 1.1 | 55.0% | 12.5 | 65.8% |
| Female | 8.3 | 59.3% | 2.3 | 76.7% | 0.8 | 40.0% | 11.4 | 60.0% |
| **Department** |  |  |  |  |  |  |  |  |
| Outpatient dept. | 8.7 | 62.1% | 2.5 | 83.3% | 0.8 | 40.0% | 12 | 63.2% |
| ART | 9.1 | 65.0% | 2.5 | 83.3% | 0.6 | 30.0% | 12.2 | 64.2% |
| IPD | 7.8 | 55.7% | 2.4 | 80.0% | 0.9 | 45.0% | 11.1 | 58.4% |
| MCH | 7.9 | 56.4% | 2.1 | 70.0% | 0.9 | 45.0% | 10.9 | 57.4% |
| TB | 11.6 | 82.9% | 2.6 | 86.7% | 1.7 | 85.0% | 15.9 | 83.7% |
| VMMC | 7.5 | 53.6% | 2.4 | 80.0% | 0.9 | 45.0% | 10.8 | 56.8% |
| Unknown | 7.5 | 53.6% | 2.7 | 90.0% | 1 | 50.0% | 11.2 | 58.9% |
| **Duration employed** |  |  |  |  |  |  |  |  |
| Less than 1 year | 8.2 | 58.6% | 2.3 | 76.7% | 0.8 | 40.0% | 11.3 | 59.5% |
| Between 1-3 years | 8.8 | 62.9% | 2.5 | 83.3% | 0.9 | 45.0% | 12.2 | 64.2% |
| Between 3-5 years | 8.3 | 59.3% | 2.4 | 80.0% | 1.1 | 55.0% | 11.8 | 62.1% |
| Between 5-10 years | 7.7 | 55.0% | 2.4 | 80.0% | 1.1 | 55.0% | 11.2 | 58.9% |
| More than 10 years | 8.7 | 62.1% | 2.3 | 76.7% | 1.1 | 55.0% | 12.1 | 63.7% |
| Unknown | 8.4 | 60.0% | 2.5 | 83.3% | 0.9 | 45.0% | 11.8 | 62.1% |
| **Ever trained on TB** |  |  |  |  |  |  |  |  |
| No | 7.9 | 56.4% | 2.3 | 76.7% | 0.8 | 40.0% | 11 | 57.9% |
| Yes | 9.1 | 65.0% | 2.5 | 83.3% | 1 | 50.0% | 12.6 | 66.3% |
| Unknown | 7.8 | 55.7% | 2.5 | 83.3% | 0.9 | 45.0% | 11.2 | 58.9% |
| **Childhood TB** |  |  |  |  |  |  |  |  |
| No | 8.8 | 62.9% | 2.5 | 83.3% | 0.9 | 45.0% | 12.2 | 64.2% |
| Yes | 9.8 | 70.0% | 2.4 | 80.0% | 1.1 | 55.0% | 13.3 | 70.0% |

In terms of departmental differences, the TB department staff scored the highest KAP (11.6 (82.9%), 2.6 (86.7%), and 1.7 (85%)), whereas the staff from the other departments scored consistently low for knowledge and practice but higher on attitudes. For instance, the KAP for OPDs was (8.7 (62.1%), 2.5 (83.3%), and 0.8 (40%)), with a high score for attitudes and a low score for knowledge and practices.

The HCWs who had been employed longer than three years scored the highest in practice (1.1 (55%)), followed by those employed between one and three years (0.9 (45%)) and those employed less than one year (0.8 (40%)). The scores for knowledge and attitudes ranged between 2.3 (76.7%) and 2.5 (83.3%) for attitude and between 8.3 (59.3%) and 8.8 (62.9%) for knowledge, except for the 5-10 years disaggregate, which scored 7.7 (55.0%).

Those who received childhood TB training performed better than those who did not. Those who had previously received childhood TB training had a KAP of 9.8 (70%), 2.4 (80%), and 1.1 (55%) compared to KAP of 8.8 (62.9%), 2.5 (83.3%), and 0.9 (45.0%) for those not previously trained.

The Table 7: Descriptive variables of HCWs KAP in the supplementary data summarizes the characteristics of the HCWs. The largest number of HCWs that took part in the study were nurses (168 (70.3%)) followed by clinical officers (53(22.2%)), then medical doctors (17(7.1%)) and (1(0.4%)) HCW did not indicate their profession.

**Table 7:** Descriptive variables of HCWs KAP

| **Summarization of qualitative characteristics** |  |
| --- | --- |
| **Variable (n=238)** | **Freq (%)** |
| **Profession** |  |
| Nurse | 168(70.3) |
| Clinical officer | 53(22.2) |
| Medical Doctor | 17 (7.1) |
| Unknown | 1 (0.4) |
| **Childhood TB closest feeling** |  |
| Compassionate and desire to help | 231 (96.7) |
| Compassionate but stay away | 1 (0.4) |
| It's their problem, can't get TB | 0 |
| I fear them, they may infect me | 1 (0.4) |
| No particular feeling | 1 (0.4) |
| Other | 1 (0.4) |
| Unknown | 4 (1.7) |
| **Reaction to work at TB corner** |  |
| Refuse | 2 (0.8) |
| Don't mind | 138 (57.7) |
| Scared | 9 (3.8) |
| Angry | 2 (0.8) |
| Happy | 77 (32.2) |
| Other | 7 (2.9) |
| Unknown | 4 (1.7) |
| **Biggest fear about TPT** |  |
| Pill burden | 69 (28.9) |
| Promotes DR-TB | 42 (17.6) |
| No benefit in high burden setting | 3 (1.3) |
| Side effects | 109 (45.6) |
| Other | 5 (2.1) |
| Unknown | 11 (4.6) |
| **How often interact with TB symptomatic children** |  |
| Everyday | 51 (21.3) |
| More than once a week | 42 (17.6) |
| At least once a week | 49 (20.5) |
| Once a month | 48 (20.1) |
| Never | 41 (17.2) |
| Unknown | 8 (3.4) |
| **Often relied on test** |  |
| Sputum microscopy | 39 (16.3) |
| Gene Xpert | 114 (47.7) |
| Chest X-ray | 47 (19.7) |
| LAM | 12 (5.0) |
| Culture and DST | 12 (5.0) |
| Other | 2 (0.8) |
| N/A | 1 (0.4) |
| Unknown | 12 (5.0) |
| **Feel well informed about TB** |  |
| No | 128 (53.6) |
| Yes | 103 (43.1) |
| Unknown | 8 (3.4) |
| **Wish to get more information about TB** |  |
| No | 0 |
| Yes | 234 (97.9) |
| Unknown | 5 (2.1) |

With regards diagnosis, 47.7 % of the HCWs indicated they rely on GeneXpert for TB diagnosis whereas 16.3%, 19.7% and 5.0% relied on Microscopy, Chest Xray and LAM respectively. Only 2 (0.8%) health care workers specified they did not know which test to use.

Majority of the healthcare workers 231 (96.7%) indicated feeling compassionate and desire to help children with TB whereas 1(0.4%) felt indifferent, another 1(0.4%) fear being infected by them and 1(0.4%) indicated feeling compassionate but tends to stay away from childhood TB patients.

When asked about their reaction if asked to work with Childhood TB, over fifty percent (138(57.7%)) indicated they didn’t mind, 77 (32.2%) responded that they’d be happy to work with children who have TB, 2 (0.8%) responded that they would refuse and wouldn’t be happy.

When asked about their biggest fear on TPT, most (109(45.6%)) of the HCWs specified the side effects as the reason. A big number of the HCWs mentioned the pill burden as a reason 69(28.9%). The HCWs that thought TPT had no benefits in high burden setting were 3(1.3%).

Finally, the Table 8: HCW Scores in the supplementary data shows the HCW variables association with the KAP score. Among the variables, the significant association was found between KAP scores and Sex, Department, Trained. Males had a higher pass rate than females although there were more females enrolled in the study. Among the various departments, all staff 4 (100%) from the TB clinic passed the KAP while the lowest pass rate came from the MCH department at 22 (57.9%) followed by IPD 48 (64.9%) and ART 23 (85.2%).

**Table 8:**
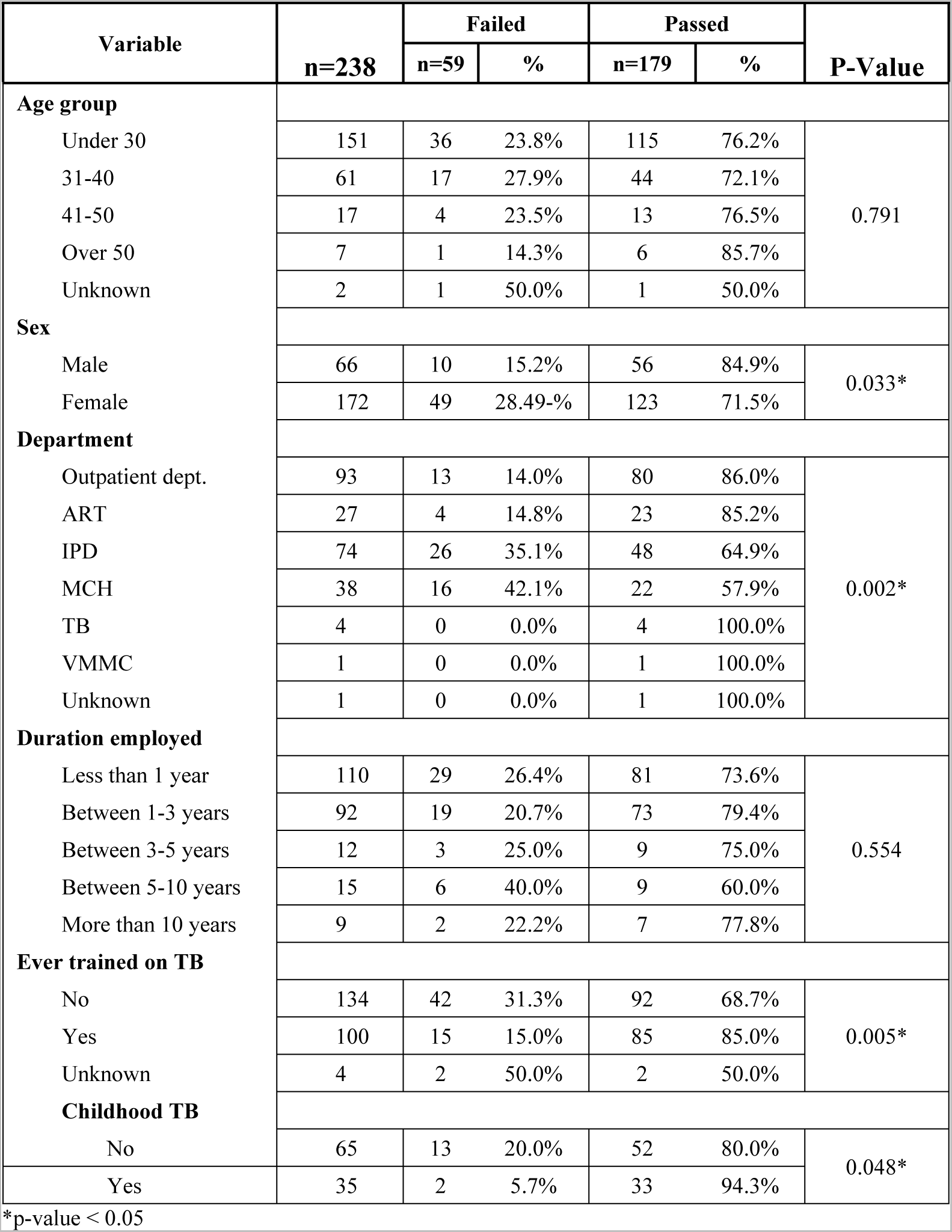
HCW Scores

| Variable | n=238 | Failed |  | Passed |  | P-Value |
| --- | --- | --- | --- | --- | --- | --- |
|  |  | n=59 | % | n=179 | % |  |
| Age group |  |  |  |  |  |  |
| Under 30 | 151 | 36 | 23.8% | 115 | 76.2% | 0.791 |
| 31-40 | 61 | 17 | 27.9% | 44 | 72.1% |  |
| 41-50 | 17 | 4 | 23.5% | 13 | 76.5% |  |
| Over 50 | 7 | 1 | 14.3% | 6 | 85.7% |  |
| Unknown | 2 | 1 | 50.0% | 1 | 50.0% |  |
| Sex |  |  |  |  |  |  |
| Male | 66 | 10 | 15.2% | 56 | 84.9% | 0.033* |
| Female | 172 | 49 | 28.49-% | 123 | 71.5% |  |
| Department |  |  |  |  |  |  |
| Outpatient dept. | 93 | 13 | 14.0% | 80 | 86.0% | 0.002* |
| ART | 27 | 4 | 14.8% | 23 | 85.2% |  |
| IPD | 74 | 26 | 35.1% | 48 | 64.9% |  |
| MCH | 38 | 16 | 42.1% | 22 | 57.9% |  |
| TB | 4 | 0 | 0.0% | 4 | 100.0% |  |
| VMMC | 1 | 0 | 0.0% | 1 | 100.0% |  |
| Unknown | 1 | 0 | 0.0% | 1 | 100.0% |  |
| Duration employed |  |  |  |  |  |  |
| Less than 1 year | 110 | 29 | 26.4% | 81 | 73.6% | 0.554 |
| Between 1-3 years | 92 | 19 | 20.7% | 73 | 79.4% |  |
| Between 3-5 years | 12 | 3 | 25.0% | 9 | 75.0% |  |
| Between 5-10 years | 15 | 6 | 40.0% | 9 | 60.0% |  |
| More than 10 years | 9 | 2 | 22.2% | 7 | 77.8% |  |
| Ever trained on TB |  |  |  |  |  |  |
| No | 134 | 42 | 31.3% | 92 | 68.7% | 0.005* |
| Yes | 100 | 15 | 15.0% | 85 | 85.0% |  |
| Unknown | 4 | 2 | 50.0% | 2 | 50.0% |  |
| Childhood TB |  |  |  |  |  |  |
| No | 65 | 13 | 20.0% | 52 | 80.0% | 0.048* |
| Yes | 35 | 2 | 5.7% | 33 | 94.3% |  |

\*p-value < 0.05

Expectedly, those who had received prior TB training had a higher pass rate 85 (85%) than those who had not 92 (68.7%) with those who received childhood TB training having even a higher pass rate of 33 (94.3%) compared to those who did not 52 (80%). Furthermore, the study shows that majority of the study participants passed 179 (75.2%) the KAP score out of the 238 total study participants.

## Discussion

A characterization of the HCW was done based on several factors. A substantial proportion had been in employment for less than 3 years and less than 50% reported ever being trained on TB and only 35% of those trained reported being trained on childhood TB. The low proportion reporting being trained on childhood TB could in part explain the low case detection of childhood TB that has been observed; Zambia has a huge gap in childhood TB case detection.

There is a considerable risk of infection for healthcare professionals, particularly nurses. Many infections can be avoided by following infection control procedures. (14, 17) This is especially key for HCWs working in a high burden TB environment because it is a highly infectious airborne disease. Our study found that a small proportion of HCW (29% of those trained) overall had previously been trained on infection control. Low infection control awareness among HCW is concerning because healthcare institutions can be prime locations for the transmission of tuberculosis (18).

The study found a significant association between TB infection control knowledge and the department where a HCW works. Staff from OPD, ART, and MCH had less understanding of TB infection control than those who did, and there were more individuals who did not respond to TB infection control questions than those who did. Despite the small number of participants from the TB clinic, health care professionals in the TB department had superior knowledge and attitudes towards TB expectedly. Healthcare workers’ understanding of routine actions that can lead to TB transmission was limited to coughing.

Majority of HCW from ART department showed poor Infection Control knowledge. This is of particular concern in view of the high-risk patients that are seen in HIV clinics. Poor infection control knowledge means that there could be a lot of nosocomial transmission in these settings. Repeated and targeted infection control training should be done frequently to improve the knowledge of staff in all departments.

Knowledge of extrapulmonary tuberculosis (ETB) and other routes of TB transmission (sneezing, singing, and laughing) should be improved. This will help improve tuberculosis Case finding and management of patients. The identification of novel markers for high and low risk persons in terms of TB development and treatment adequacy would enable for evidence-based decisions about who to treat, how to treat, and for how long to treat for both prevention and cure.

While HCWs from other departments (OPD, ART, and MCH) showed a good degree of knowledge of TB symptoms and a desire to participate in TB activities, there is still a need to increase the engagement of HCWs from other departments (OPD, ART, and MCH) in TB evaluation. In addition, the TB program must invest more in educating HCWs outside of the TB clinic. This is similar to recommendations made by An et al “*further investments and efforts should be considered to translate knowledge into action to ensure that children with presumptive TB will be duly screened, detected, and treated in low TB case detection ODs to improve childhood TB detection in Cambodia*” (19)

Although referring children to the TB clinic is the standard practice for HCWs who are not mainly in charge of TB treatment, which 73.1% of HCWs do, 55.5% of them claimed they also help collect sputum samples before referring the kids to further TB services. Therefore, it is essential that these healthcare workers receive infection control training because they are likely to contract TB themselves.

The study showed that there are varied knowledge levels based on the gender, department and training history of the healthcare workers. Consequently, there is need to improve the TB knowledge, attitudes and practices of the female health care workers as they make up majority of the staff in TB diagnosis and treatment. Additionally, there is need to include staff from other departments that are not directly responsible for TB treatment as trainings about TB are being conducted. This was shown in the study where all HCWs from the TB clinic 4 (100%) passed the KAP while departments like IPD and ART only scored a pass rate of 22 (57.9%) and 48 (64.9%) respectively. However, the OPD had a good score at 80 (86.0%) indicating that there is good KAP in the main departments involved in TB diagnosis with the study further showing that staff with prior TB specific training have significantly high KAP scores at 33 (94.3%) compared to those who did not have prior TB training 52 (80.0%).

### Strengths of the study

A total of 238 HCWs were targeted and all of them were enrolled into the study, bringing the study to 100% response rate and all subjects were included in the analysis. Our study was a random sample of the population and we used the recommended KAP survey adapted from the sample questionnaire on KAP studies on TB by WHO and StopTBPartnership (20).

### Limitations

The study was conducted in Lusaka Urban communities and may not be representative of the population of Zambia although it does share similar features with other urban and peri-urban communities., and the knowledge attitudes and practices differ between urban and rural settings. Future studies should look at recruiting participants from multiple settings. We might have experienced some unintended selection bias in the health care workers enrolled in the study as some of the departments had shift workers that may not have been present at the time the questionnaires were being administered.

The other limitation is that the analysis was not done to compare the setting before intervention and after.

## Declarations

**Ethics approval** was obtained from the University of Zambia Biomedical and Research Ethics Committee (IRB Reference number 635-2020).

**Availability of Data and materials** upon request from as per Data sharing policies of the institution.

## CONFLICT OF INTEREST

We the authors declare that we are free of any conflict of interest.

## FUNDING

The Stop TB Partnership/ TB REACH mechanism with funding support from the Government of Canada (TB REACH wave 7)

## AUTHORS CONTRIBUTIONS

Study Conceptualization: Mary Kagujje, Data collection Brian Shuma, Lophina Chilukutu, Sarah Nyangu

Data curation: data analysis Minyoi Maimbolwa, Paul Kaumba

Script write-up: data interpretation and revisions were done by Paul Kaumba, Mary Kagujje, Sarah Nyangu, Monde Muyoyeta, Nsala Sanjase,

All authors have reviewed the manuscript and agree to publish it.

## Data Availability

ll relevant data are within the manuscript and its Supporting Information files.

## ACKNOWLEDGEMENTS

The authors would like to thank the following for their support and contributions:

The staff from the chest clinic/TB corners at Chawama first level hospital and Kanyama first level hospital.

TB treatment supporters in all health facilities.

The Lusaka district health office for creating a good working environment for the study team in the health facilities.

## Notes

### Competing Interest Statement

The authors have declared no competing interest.

### Funding Statement

Dr. Monde Muyoyeta received the a grant through the Stop TB Partnership/ TB REACH mechanism with funding support from the Government of Canada (TB REACH wave 7) which facilitated the collection of the data for this study. This funding was obtained under The Centre for Infectious Diseases Research in Zambia (CIDRZ). Grant Number: STBP/TBREACH/GSA/W7-7426 Funders site: https://stoptb.org/global/awards/tbreach/wave7.asp The funders had no role in study design, data collection and analysis, decision to publish, or preparation of the manuscript.

### Author Declarations

Ethics approval was obtained from the University of Zambia Biomedical and Research Ethics Committee (IRB Reference number 635-2020).

## References

1. WHO. GLOBAL TB REPORT. 2021.

2. Dodd PJ, Yuen CM, Sismanidis C, Seddon JA, Jenkins HE. The global burden of tuberculosis mortality in children: a mathematical modelling study. The Lancet Global health. 2017;5(9):e898–e906.

3. Partnership S. 2020.

4. Reuter A, Hughes J, Furin J. Challenges and controversies in childhood tuberculosis. The Lancet. 2019;394(10202):967–78.

5. Kissou SA, Millogo JDC, Nikiema Z, Birba E, Cessouma R, Sanogo B, et al. Childhood Tuberculosis in Sub-saharan Africa. International Journal of Pediatrics & Neonatal Care. 2018;4.

6. Bashorun AO, Linda C, Omoleke S, Kendall L, Donkor SD, Kinteh M-A, et al. Knowledge, attitude and practice towards tuberculosis in Gambia: a nation-wide cross-sectional survey. BMC public health. 2020;20(1):1–13.

7. Alotaibi B, Yassin Y, Mushi A, Maashi F, Thomas A, Mohamed G, et al. Tuberculosis knowledge, attitude and practice among healthcare workers during the 2016 Hajj. PLoS One. 2019;14(1):e0210913.

8. Farhanah AW, Sarimah A, Jafri Malin A, Hasnan J, Siti Suraiya MN, Wan Mohd Zahiruddin Wan M, et al. Updates on knowledge, attitude and preventive practices on tuberculosis among healthcare workers. Malaysian Journal of Medical Sciences. 2016;23(6):25–34.

9. Sima BT, Belachew T, Abebe F. Health care providers’ knowledge, attitude and perceived stigma regarding tuberculosis in a pastoralist community in Ethiopia: a cross-sectional study. BMC health services research. 2019;19(1):1–11.

10. Shrestha A, Bhattarai D, Thapa B, Basel P, Wagle RR. Health care workers’ knowledge, attitudes and practices on tuberculosis infection control, Nepal. BMC Infectious Diseases. 2017;17(1):1–7.

11. Noé A, Ribeiro RM, Anselmo R, Maixenchs M, Sitole L, Munguambe K, et al. Knowledge, attitudes and practices regarding tuberculosis care among health workers in Southern Mozambique. BMC pulmonary medicine. 2017;17(1):1–7.

12. Minnery M, Contreras C, Pérez R, Solórzano N, Tintaya K, Jimenez J, et al. A cross sectional study of knowledge and attitudes towards tuberculosis amongst front-line tuberculosis personnel in high burden areas of Lima, Peru. PLoS One. 2013;8(9):e75698.

13. Marais B, Graham S, Cotton M, Beyers N. Diagnostic and management challenges for childhood tuberculosis in the era of HIV. The Journal of infectious diseases. 2007;196(Supplement_1):S76–S85.

14. WHO. Guidelines for the Management of Latent Tuberculosis Infection. 2020.

15. Keïta M, Camara AY, Traoré F, Camara ME, Kpanamou A, Camara S, et al. Impact of infection prevention and control training on health facilities during the Ebola virus disease outbreak in Guinea. BMC Public Health. 2018;18(1):1–7.

16. Organization WH. Advocacy, communication and social mobilization for TB control: a guide to developing knowledge, attitude and practice surveys. World Health Organization; 2008. Report No.: 9241596171.

17. Joshi R, Reingold AL, Menzies D, Pai M. Tuberculosis among Health-Care Workers in Low- and Middle-Income Countries: A Systematic Review. PLOS Medicine. 2006;3(12):e494.

18. Xie Z, Zhou N, Chi Y, Huang G, Wang J, Gao H, et al. Nosocomial tuberculosis transmission from 2006 to 2018 in Beijing Chest Hospital, China. Antimicrobial resistance and infection control. 2020;9(1):165.

19. An Y, Teo AKJ, Huot CY, Tieng S, Khun KE, Pheng SH, et al. Knowledge, attitude, and practices regarding childhood tuberculosis detection and management among health care providers in Cambodia: a cross-sectional study. BMC Infectious Diseases. 2022;22(1)yyyy:1–11.

20. WHO. A guide to developing knowledge, attitude and practice surveys. 2008.

